# Sharing ventilators in the Covid-19 pandemics. A bench study

**DOI:** 10.1101/2020.11.02.20224774

**Authors:** Claude Guérin, Martin Cour, Neven Stevic, Florian Degivry, Erwan L’Her, Bruno Louis, Laurent Argaud

## Abstract

COVID-19 pandemics sets the healthcare system to a shortage of ventilators. We aimed at assessing tidal volume (V_T_) delivery and air recirculation during expiration when one ventilator is divided into 2 patients. The study was performed in a research laboratory in a medical ICU of a University hospital. An ICU-dedicated (V500) and a lower-level ventilator (Elisée 350) were attached to two test-lungs (QuickLung) through a dedicated flow-splitter. A 50 mL/cmH_2_O Compliance (C) and 5 cmH_2_O/L/s Resistance (R) were set in both A and B lungs (step1), C50R20 in A / C20R20 in B (step 2), C20R20 in A / C10R20 in B (step 3), and C50R20 in A / C20R5 in B (step 4). Each ventilator was set in volume and pressure control mode to deliver 0.8L V_T_. We assessed V_T_ from a pneumotachograph placed immediately before each lung, rebreathed volume, and expiratory resistance (circuit and valve). Values are median (1^st^-3^rd^ quartiles) and compared between ventilators by non-parametric tests. Between Elisée 350 and V500 in volume control V_T_ in A/B patients were 0.381/0.387 vs. 0.412/0.433L in step 1, 0.501/0.270 vs. 0.492/0.370L in step 2, 0.509/0.237 vs. 0.496/0.332L in step 3, and 0.496/0.281 vs. 0.480/0.329L in step 4. In pressure control the corresponding values were 0.373/0.336 vs. 0.430/0.414L, 0.416/0.185/0.322/0.234L, 0.193/0.108 vs. 0.176/0.092L and 0.422/0.201 vs. 0.481/0.329L, respectively (P<0.001 between ventilators at each step for each volume). Rebreathed air volume ranged between 0.7 to 37.8 ml and negatively correlated with expiratory resistance in steps 2 and 3. The lower-level ventilator performed closely to the ICU-dedicated ventilator. Due to dependence of V_T_ to C pressure control should be used to maintain adequate V_T_ at least in one patient when C and/or R changes abruptly and monitoring of V_T_ should be done carefully. Increasing expiratory resistance should reduce rebreathed volume.

## Introduction

During the COVID-19 pandemic, a risk of a shortage of ICU ventilators was claimed very early [1]. As the poliomyelitis pandemic prompted the caregivers to discover tracheotomy, iron lung, and mechanical ventilation, the current COVID-19 pandemic prompted innovative solutions [2]. They include ventilator multipliers, portable and open-source designs of ventilators [3], and frugal ventilators [4]. Sharing ventilation provides ventilatory support to two or more patients with the same ventilator [5]. This approach raised ethical issues [6] due to the many technical problems to solve from sharing the same ventilator with patients with different respiratory mechanics and, hence different requirements [7]. For multiplex ventilation, with no means of independently controlling positive end-expiratory pressure (PEEP) and tidal volume (V_T_), patients sharing the same ventilator should have respiratory mechanics as similar as possible. In this case, in volume control ventilation and pressure control ventilation mode, each patient is expected to receive half of the set V_T_. Any decrease in compliance and/or increase in resistance in one patient will decrease V_T_ in each mode [8, 9]. For the other patient with unchanged compliance and resistance, V_T_ will depend on a part on mechanical ventilation mode. The degree to which differences in mechanics cause differences in tidal volume depends both on how the mechanics differ and on the mode of ventilation (ie volume versus pressure control), but in general, changes in one patient cause changes in the other patient regardless of the model [8]. Moreover, air may recirculate during inspiration and expiration from one patient to the other, with the risk of CO_2_ retention and cross-transmission of infection. The patient with the shortest inspiratory time constant, i.e. with the lowest product of resistance by compliance, breathed out faster (earlier) while the other was still filling in. However, the role of expiratory resistance (circuit and ventilator valve) on rebreathed air has not been previously addressed. Nevertheless, the feasibility and safety of ventilator sharing have been reported recently in a few patients highly selected, deeply monitored, and for a few hours [10-12].

Because ventilator sharing is still experimental and not completely investigated we designed a bench study where 2 or 3 patients with different respiratory mechanics were attached to the same ventilator. We compared a high-performance ICU ventilator and a ventilator used for patient transportation. We assessed V_T_ delivery, rebreathed volume, and expiratory resistance.

## Methods

Two ventilators were tested: the Elisée 350 (ResMed, Saint-Priest, France) a turbine-driven ventilator used for patient transportation, and in stepdown-units and the V500 ICU ventilator (Drager, Lubeck, Germany). They were attached to two QuickLung tests (IngMar Medical, Inc., Pittsburgh, PA) equipped with resistance (R) of 5, 20, and 50 cmH_2_O/L/s and compliance (C) of 10, 20, and 50 mL/cmH_2_O in the first part of the experiment. In the second part a third lung test (SelfTestLung, Draeger, Lubeck, Germany) of 10 mL/cmH_2_O C and 20 cmH_2_O/L/s ± was added (Table 1).

**Table 1.** Study design.

| Parts | Steps | Patient A | Patient B | Patient C<br>SelfTestLung |
| --- | --- | --- | --- | --- |

|  |  | QuickTest<br>lung | QuickTest<br>lung |  |
| --- | --- | --- | --- | --- |
| I. Two Lungs design | 1 | C50-R5 | C50-R5 | none |
|  | 2 | C50-R20 | C20-R20 | none |
|  | 3 | C20-R20 | C10-R20 | none |
|  | 4 | C50-R20 | C20-R5 | none |
| II. Three Lungs design | 1 | C50-R5 | C50-R5 | C10-R20 |
|  | 2 | C50-R20 | C20-R20 | C10-R20 |
|  | 3 | C20-R20 | C10-R20 | C10-R20 |
|  | 4 | C50-R20 | C20-R5 | C10-R20 |

C: compliance in ml/cmH<sub>2</sub>O, R: resistance in cmH<sub>2</sub>O/L/s

Besides, standard double-limb ventilator circuit of 22 mmID (Intersurgical, Fontenay sous bois, France), high-efficiency particulate air filter (HEPA Isogard, Gibeck, Indianapolis, IN) in front of each test lung and specific flow-splitters (MICHELIN Molding Solutions, Michelin, Clermont-Ferrand, France) were used (Figure 1). Airflow was measured by pneumotachographs (Hamilton, Sidam, Mirandola, Italy) and airway pressure (Paw) by pressure transducers (Gabarith PMSET 1DT-XX, Becton Dickinson, Singapore).

**Fig 1.**
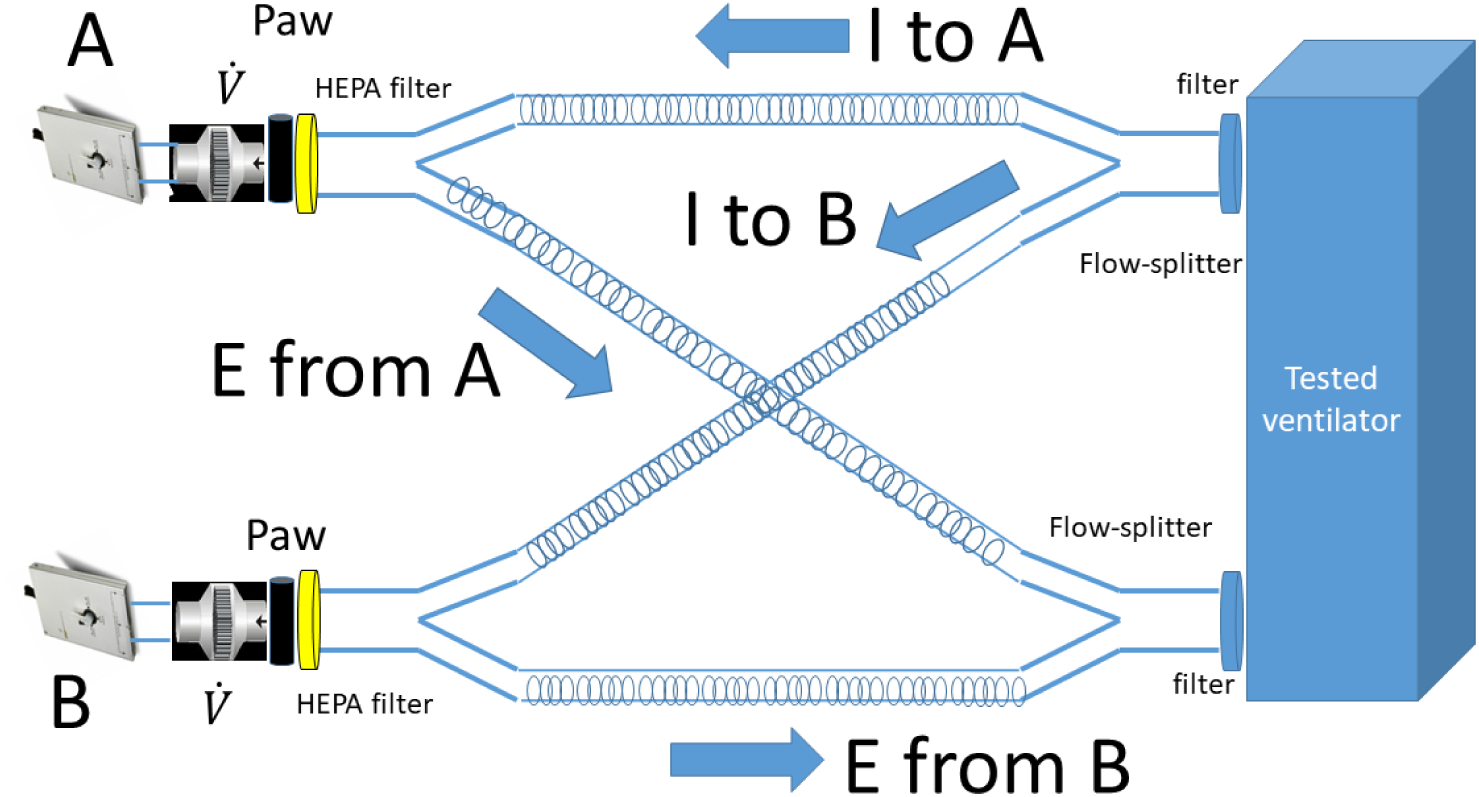
Experimental set-up with two lungs A and B. Paw airway pressure, I inspiration, E expiration, HEPA High-Efficiency Particulate air filter, 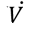 flow.

Two designs were used for each ventilator in both volume and pressure control ventilation (Table 1).

In the first part, the two QuickLung models were arranged in parallel (Fig 1) and 4 steps performed. In step 1a similar C50R5 was applied to both patients, to replicate type L COVID-19-related acute respiratory distress syndrome (ARDS) [13]. The 3 other steps had contrasted time constants: step 2 (C50R20 vs. C20R20), step 3 (C20R20 vs. C10R20), and step 4 (C50R20 vs. C20R5) (Table 1). In volume control, V_T_ was set to 0.800 L to deliver 0.4 L to each patient, i.e. 6 mL/kg V_T_ for the average predicted body weight in patients with ARDS in the Lung Safe study [14], PEEP to 15 cmH_2_O, a value chosen to stretch the ventilator, and inspired oxygen fraction (FIO_2_) to 21%. In pressure control, ventilator was set to get a V_T_ of 0.8 L in step1. This setting was kept for the other steps. The other ventilator settings are displayed in Table 2. The values of C we have selected were the options allowed by the lung model we used. In COVID-19 related ARDS, C spread out from 20 to 90 ml/cmH2O, with 50 ml/cmH2O the most frequent value [15]. Further studies reported values of C in the range of 26-65 ml/cmH2O [16-18].

**Table 2.**
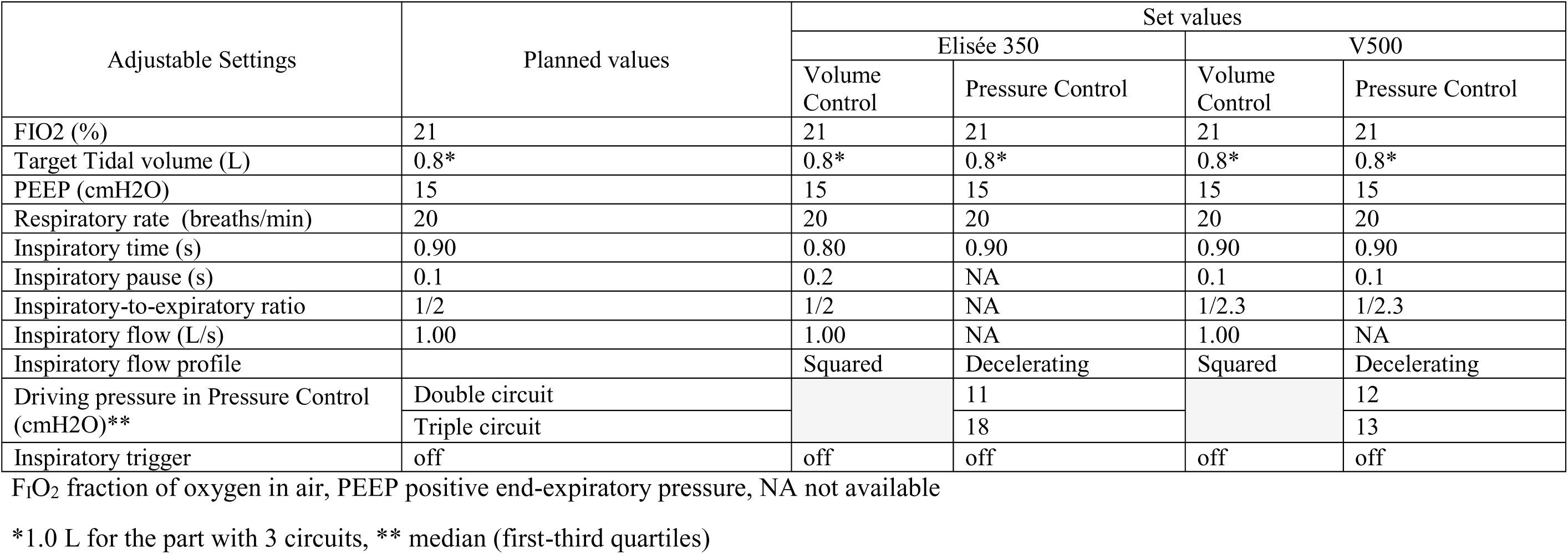
Settings of the ventilators during the bench study.

| Adjustable Settings | Planned values | Set values |  |  |  |
| --- | --- | --- | --- | --- | --- |
|  |  | Elisée 350 |  | V500 |  |
|  |  | Volume Control | Pressure Control | Volume Control | Pressure Control |
| FIO2 (%) | 21 | 21 | 21 | 21 | 21 |
| Target Tidal volume (L) | 0.8* | 0.8* | 0.8* | 0.8* | 0.8* |
| PEEP (cmH2O) | 15 | 15 | 15 | 15 | 15 |
| Respiratory rate (breaths/min) | 20 | 20 | 20 | 20 | 20 |
| Inspiratory time (s) | 0.90 | 0.80 | 0.90 | 0.90 | 0.90 |
| Inspiratory pause (s) | 0.1 | 0.2 | NA | 0.1 | 0.1 |
| Inspiratory-to-expiratory ratio | 1/2 | 1/2 | NA | 1/2.3 | 1/2.3 |
| Inspiratory flow (L/s) | 1.00 | 1.00 | NA | 1.00 | NA |
| Inspiratory flow profile |  | Squared | Decelerating | Squared | Decelerating |
| Driving pressure in Pressure Control (cmH2O)** | Double circuit |  | 11 |  | 12 |
|  | Triple circuit |  | 18 |  | 13 |
| Inspiratory trigger | off | off | off | off | off |

F<sub>I</sub>O<sub>2</sub> fraction of oxygen in air, PEEP positive end-expiratory pressure, NA not available
\*1.0 L for the part with 3 circuits, \*\* median (first-third quartiles)

In pressure control, the pressure was adjusted to deliver a V_T_ of 0.800 L and settings were unaltered because the steps of the experiment (Table 1) were thought to reflect the time course of asymmetrical disturbance that may occur between patients attached to the same ventilator, i.e. sudden loss of lung volume in one patient (pneumothorax, atelectasis), and hence the ventilator has no reason to be set differently before the situation occurs. Such a double circuit design is also suitable at the time of patient selection for parallel ventilation. The safety guard is to share the single ventilator between patients with respiratory mechanics as close as possible [7, 19]. We wanted to explore how much V_T_ would depart from each patient to the other when the respiratory mechanics markedly differ between them. In the second design, a third lung with a fixed C20-R20 was added in parallel to the previous C-R (Table 1) of the two QuickLung models and V_T_ set to 1.0 L. This part of the study aimed at stretching the asymmetry between patients. Our choice of C-R was in line with the standards [20].

Flow and Paw analog signals were sent to a datalogger (Biopac MP150, Biopac Inc., Goletta, CA). Mechanical ventilation was stabilized for one minute and, then signals were collected for one minute at a 200 Hz sampling rate.

### Data analysis

Collected data were analyzed off-line by using in-house software specifically developed for the present study (Matlab R2019b, MathWorks, Inc.). V_T_ was obtained by integration of the flow signal over the inflation time in each lung, which can be different between each lung (see below) and different from the machine inflation time Taking care of this an important in design, like our present one, that accommodates time constants differences between lungs. The rebreathed volume was computed as the amount of air that flowed from one patient to the other(s) during inspiration and expiration (Figure 2).

**Figure 2.**
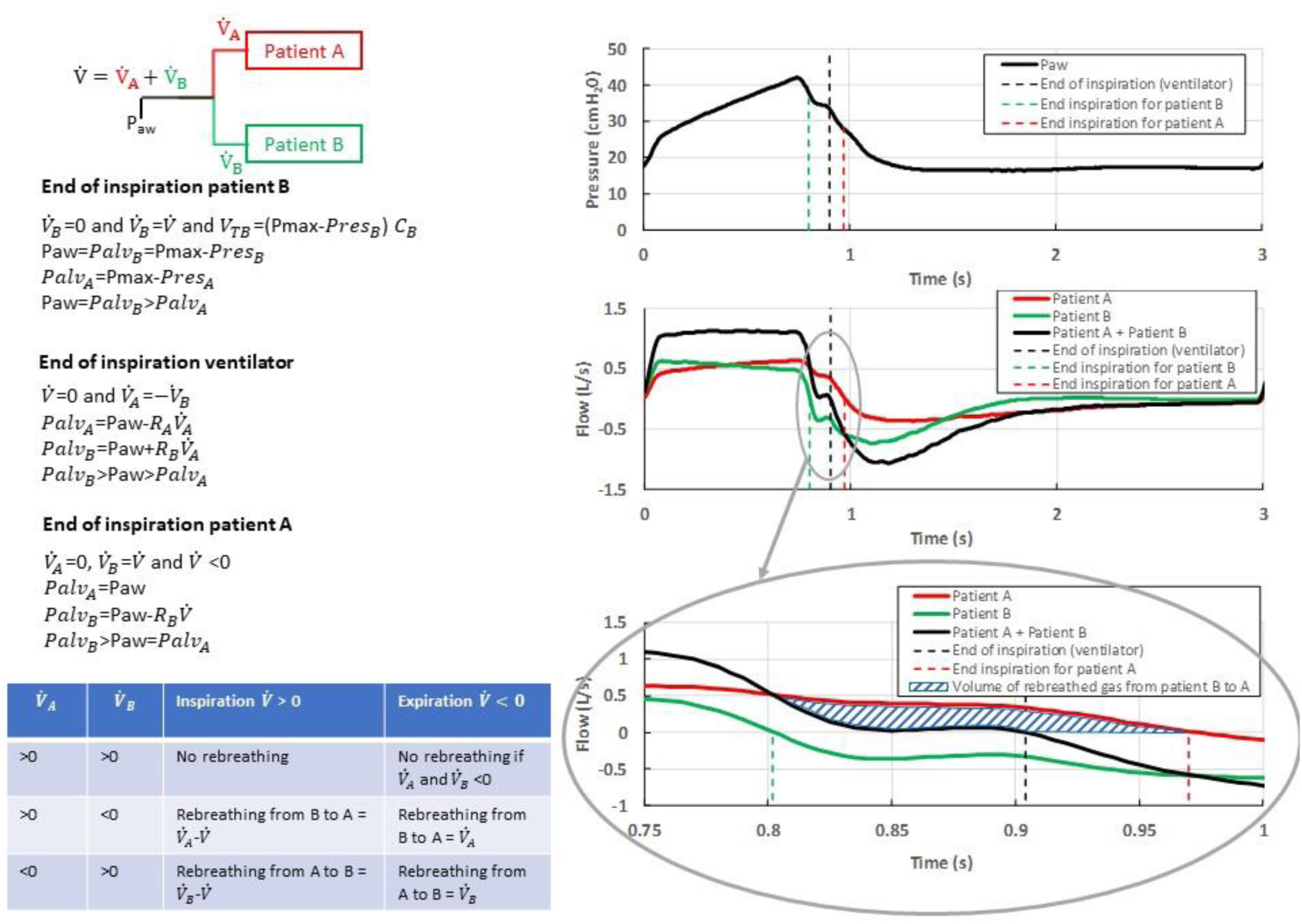
Method to measure the rebreathed volume during inspiration and expiration in case of a double circuit and two patients with uneven compliance and resistance ventilated in volume control. The schematic upper left drawing illustrates the airway pressure (Paw) and the flow 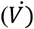coming from the ventilator. This is distributed in parallel to patient A (red) 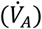 and patient B (green) 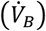. On the right part recording of Paw (top) and flow (middle) from the ventilator, patient A and patient B overtime during a breathing cycle. The vertical broken lines indicate the corresponding ends of insufflation. The lower right panel is a magnification of the end of insufflation.

The patient B with the shortest inspiratory time constant ends up inspiration earlier than patient A. At the time of patient B end of inspiration, 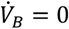, and 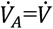. The tidal volume received by patient B is equal to the compliance of the lung B times maximal Paw (Pmax) minus resistive pressure (PresB), which equal to zero because *of* 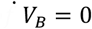. Therefore Pmax equals alveolar pressure in lung B (PalvB). At the same time since 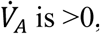, PalvA is equal to Pmax minus PresA, and hence is lower than PalvB.

At the time of the ventilator end of insufflation, 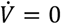, and 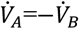. PalvA is equal to Paw minus PresA (which is the product of resistance through lung A to 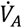 and similarly PalvB is equal to Paw plus PresB (which is the product of resistance through lung B to 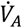). It comes that PalvB>Paw>PalvA.

At the time of patient A end of inspiration, 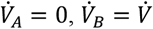, *and* 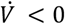. Therefore, PalvA=Paw and PalvB is equal to Paw minus PresB (which is the product of the resistance through lung B to 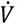). PalvB is then greater than Paw, which is equal to PalvA.

The table in the bottom left summarizes the conditions making rebreathing to or not to happen and its computation. The hatched area in the lower right graph indicates the amount of air rebreathing from patient B to patient A. A similar approach was used for pressure control (not shown).

On each breath, the instantaneous expiratory resistance was determined as the ratio of the pressure drop between Paw and atmosphere to the corresponding flow as previously described [21]. For this computation, we discarded flows lower than 0.01 L/S to avoid extreme values corresponding to the closing of the valve. Therefore, the instantaneous expiratory resistance was determined in roughly 400 instances in each breath. We used the minimal value of the instantaneous resistance in each condition.

The primary end-point was the value of V_T_ and secondary end-points were rebreathed volume and minimal instantaneous expiratory resistance. Values are presented as median (1st-3rd quartiles) and compared across ventilators for each C-R condition by using a non-parametric test. The statistically significant level was set to P-value < 0.05. The analysis was performed by using R software Version 3.5.2 (R: A Language and Environment for Statistical Computing, R Core Team, R Foundation for Statistical Computing, Vienna, Austria, 2018).

## Results

### V_T_ delivery

As expected for the step1, V_T_ was equally delivered to patients A and B in both Volume and Pressure Control modes (Figure 3). Between Elisée 350 and V500 ventilators with the double circuit, V_T_ amounted to 0.381 (0.379-0.382) L in lung A and 0.387 (0.385-0.389) L in lung B and 0.412 (0.409-0.413) L and 0.433 (0.432-0.435) L, respectively (P<0.05 between ventilators) in volume control, and 0.373 (0.372-0.375) L in lung A and 0.336 (0.336-0.338) L in lung B versus 0.430 (0.429-0.430) and 0.414 (0.413-0.416) L, respectively in pressure control (P<0.05 between ventilators). Both ventilators accurately delivered the targeted value of V_T_ within 10% boundaries, with Elisée 350 under-delivering V_T_ by 6 (5-7)% and V500 over-delivering V_T_ by 4 (2-7)%.

**Fig 3.**
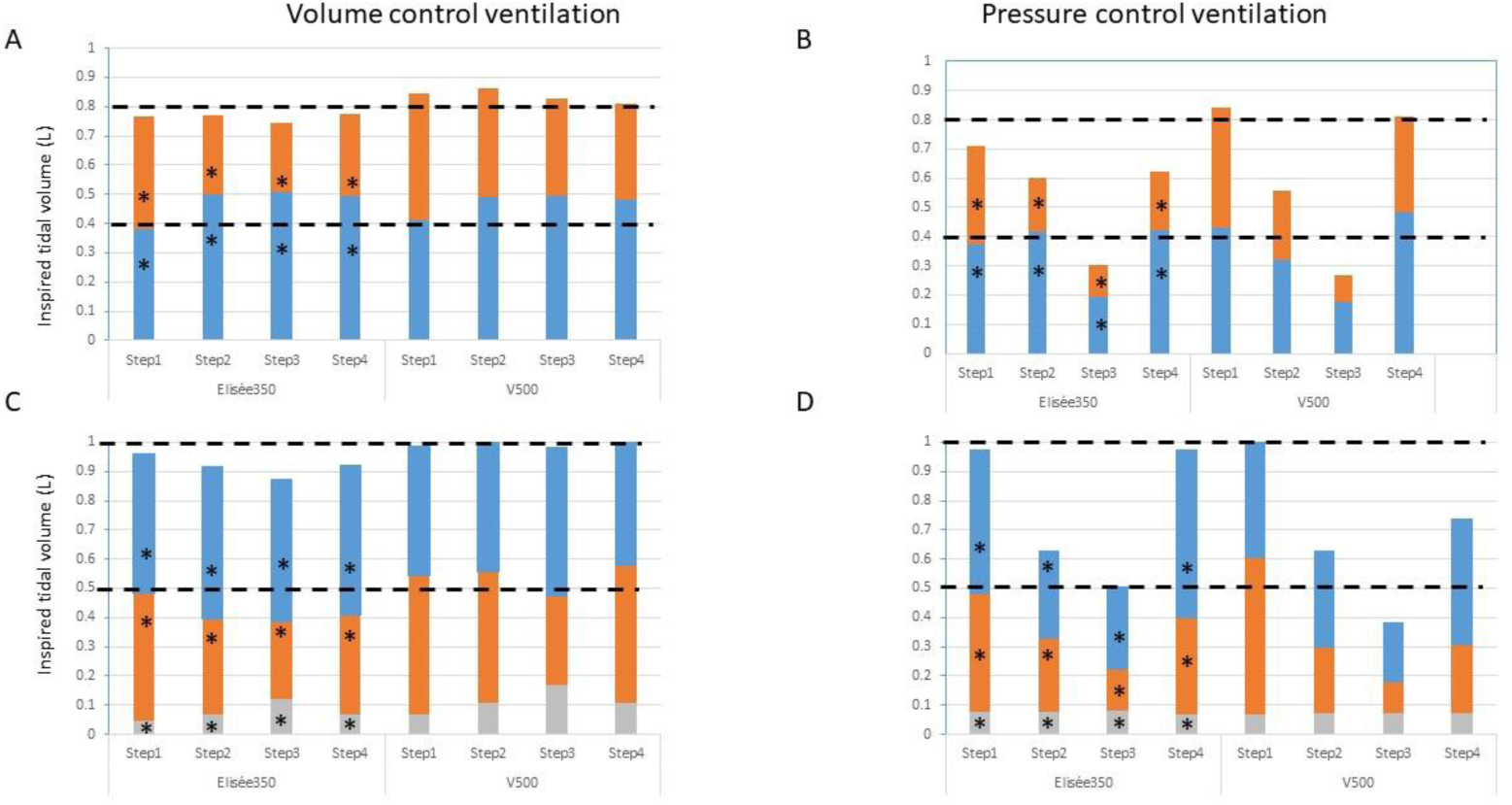
Inspired tidal volume. Stacked plots of inspired tidal volume in patient A (blue), B (orange), and C (grey) in volume and pressure control ventilation during the 4 steps for each ventilator in the design with two (panels A and B) and three patients (panels C and D). Step1 = C50-R5 for each patient, Step2 = C50-R20 in A and C20-R20 in B, Step3 = C20-R20 in A and C10-R20 in B, Step4 = C50-R20 in A and C20-R5 in B. C25-R20 for patient C at each step. Bars are median and quartiles omitted for clarity. The horizontal broken black lines indicate the target tidal volume the ventilator should deliver to each patient. *P<0.05 as compared to V500.

In step 2 C was 2.5 times greater in patient A than in B and R was similar in both of them but 4 times greater than in step 1. In volume control, V_T_ was greater in patient A than in patient B by a factor of 1.9 with Elisée 350 and 1.33 with V500 (Figure 3). It was 0.501 (0.500-0.503) L in patient A and 0.270 (0.269-0.271) L in patient B with Elisée and 0.492 (0.492-0.493) L and 0.70 (0.360-0.381) L, respectively (P<0.001 between ventilators). Therefore, V_T_ delivery was 20% greater in patient A and 33% lower in patient B than expected with Elisée, these values being of 23% and 8%, respectively, with V500. A similar figure was observed in step 3 where C was 2 times greater in patient A than in patient B but 2.5 and 5 times lower, respectively, as compared to step 1, and R was similar in both lungs and similar to in step 3 (Figure 3). The same was true for step 4 (Figure 2). Contrary to step 1, in steps 2-4 V_T_ to patient A was greater with Elisée 350 than with V500 and the opposite was true for V_T_ to lung B (Figure 3).

In pressure control with the asymmetrical design, V_T_ in a given patient changed as a result of both the overall V_T_ decrease due to the greater impedance and the difference between time constants. In step 2, V_T_ was greater in patient A than in B by a factor of 2.2 with Elisée 350 and of 1.4 with V500 (Figure 3). However, patient A accurately received the target V_T_ (+4%), whilst patient B had V_T_ reduced by 54% as compared to the target V_T_ with Elisée 350. By contrast, with V500 in patient A V_T_ was under-delivered by 20% as compared to the target V_T_ whilst patient B received V_T_ reduced by 43% from the target V_T_ (P<0.001 between ventilators). The same picture was observed in steps 3 and 4.

In the triple circuit, V_T_ delivered to patient C in volume control was very small according to its low C, with lower values with Elisée 350 than V500 (Figure 3C). In step 1 V_T_ to patient C was 0.044 (0.044-0.045) and 0.076 (0.076-0.076) L with Elisée 350 and V500, respectively, (P<0.001 between ventilators), representing 5 and 8% of whole delivered V_T_. In the asymmetrical steps 2-4, in particular with V500 V_T_ to patient C increased up to 0.134 (0.133-0.136) L in step 2, 0.176 (0.175-0.178) L in step 3, and 0.122 (0.121-0.123) L in step 4, which sets this stiff lung to the risk of overdistension. In pressure control V_T_ to patient C was 0.067 (0.067-0.067) L with Elisée 350 and 0.075 (0.074-0.077) L with V500 is step 1 (Figure 3D). However, in the asymmetrical 2-4 steps, V_T_ delivered to that patient remained stable and in line with its low C preserving it from overdistension (Figure 3D).

### Rebreathed volume

In the double circuit, the rebreathed volume ranged from 0.7 to 37.8 ml (Table 3). The direction of the significant differences between ventilators varies across conditions. The amount of rebreathed volume tended to be lower in pressure control than in volume control. The picture was essentially the same in the triple circuit. The rebreathed volume to patient C was very small, as expected from its very low C. The volume of the rebreathed volume was small as compared to the volume of the expiratory circuit (607 mL).

**Table 3.**
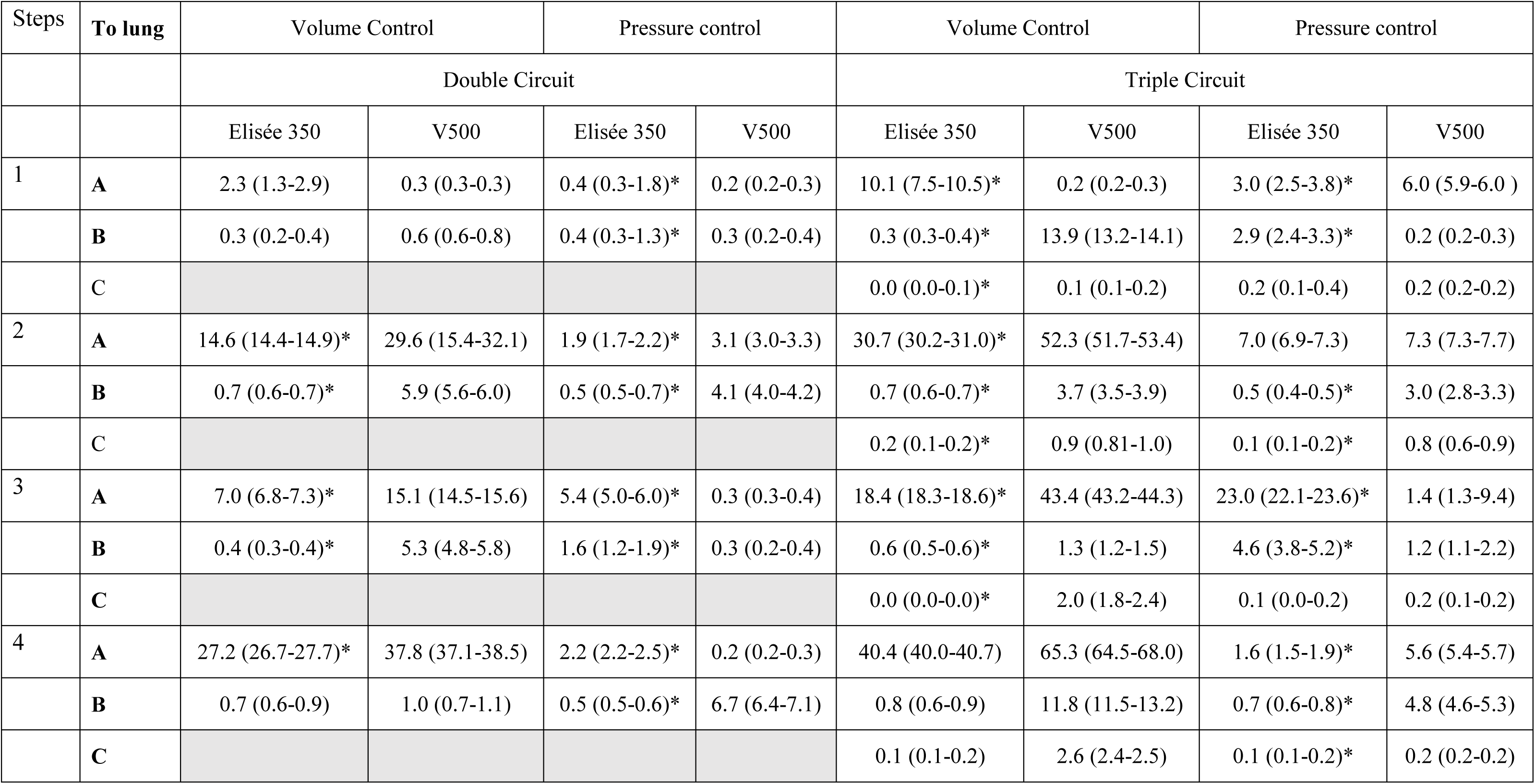
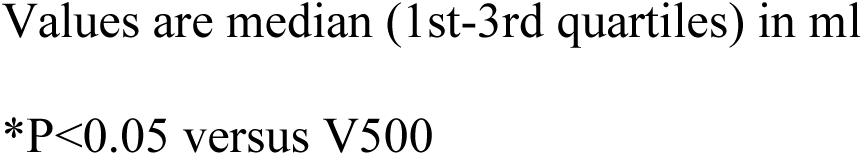
Rebreathed volume to each lung.

| Steps | To lung | Volume Control |  | Pressure control |  | Volume Control |  | Pressure control |  |
| --- | --- | --- | --- | --- | --- | --- | --- | --- | --- |
|  |  | Double Circuit |  |  |  | Triple Circuit |  |  |  |
|  |  | Elisée 350 | V500 | Elisée 350 | V500 | Elisée 350 | V500 | Elisée 350 | V500 |
| 1 | A | 2.3 (1.3-2.9) | 0.3 (0.3-0.3) | 0.4 (0.3-1.8)* | 0.2 (0.2-0.3) | 10.1 (7.5-10.5)* | 0.2 (0.2-0.3) | 3.0 (2.5-3.8)* | 6.0 (5.9-6.0 ) |
|  | B | 0.3 (0.2-0.4) | 0.6 (0.6-0.8) | 0.4 (0.3-1.3)* | 0.3 (0.2-0.4) | 0.3 (0.3-0.4)* | 13.9 (13.2-14.1) | 2.9 (2.4-3.3)* | 0.2 (0.2-0.3) |
|  | C |  |  |  |  | 0.0 (0.0-0.1)* | 0.1 (0.1-0.2) | 0.2 (0.1-0.4) | 0.2 (0.2-0.2) |
| 2 | A | 14.6 (14.4-14.9)* | 29.6 (15.4-32.1) | 1.9 (1.7-2.2)* | 3.1 (3.0-3.3) | 30.7 (30.2-31.0)* | 52.3 (51.7-53.4) | 7.0 (6.9-7.3) | 7.3 (7.3-7.7) |
|  | B | 0.7 (0.6-0.7)* | 5.9 (5.6-6.0) | 0.5 (0.5-0.7)* | 4.1 (4.0-4.2) | 0.7 (0.6-0.7)* | 3.7 (3.5-3.9) | 0.5 (0.4-0.5)* | 3.0 (2.8-3.3) |
|  | C |  |  |  |  | 0.2 (0.1-0.2)* | 0.9 (0.81-1.0) | 0.1 (0.1-0.2)* | 0.8 (0.6-0.9) |
| 3 | A | 7.0 (6.8-7.3)* | 15.1 (14.5-15.6) | 5.4 (5.0-6.0)* | 0.3 (0.3-0.4) | 18.4 (18.3-18.6)* | 43.4 (43.2-44.3) | 23.0 (22.1-23.6)* | 1.4 (1.3-9.4) |
|  | B | 0.4 (0.3-0.4)* | 5.3 (4.8-5.8) | 1.6 (1.2-1.9)* | 0.3 (0.2-0.4) | 0.6 (0.5-0.6)* | 1.3 (1.2-1.5) | 4.6 (3.8-5.2)* | 1.2 (1.1-2.2) |
|  | C |  |  |  |  | 0.0 (0.0-0.0)* | 2.0 (1.8-2.4) | 0.1 (0.0-0.2) | 0.2 (0.1-0.2) |
| 4 | A | 27.2 (26.7-27.7)* | 37.8 (37.1-38.5) | 2.2 (2.2-2.5)* | 0.2 (0.2-0.3) | 40.4 (40.0-40.7) | 65.3 (64.5-68.0) | 1.6 (1.5-1.9)* | 5.6 (5.4-5.7) |
|  | B | 0.7 (0.6-0.9) | 1.0 (0.7-1.1) | 0.5 (0.5-0.6)* | 6.7 (6.4-7.1) | 0.8 (0.6-0.9) | 11.8 (11.5-13.2) | 0.7 (0.6-0.8)* | 4.8 (4.6-5.3) |
|  | C |  |  |  |  | 0.1 (0.1-0.2) | 2.6 (2.4-2.5) | 0.1 (0.1-0.2)* | 0.2 (0.2-0.2) |

Values are median (1st-3rd quartiles) in ml
\*P<0.05 versus V500

### Expiratory resistance

The minimal expiratory resistance was different between ventilators in most instances in Figure 4. The differences between ventilators were statistically significant in every comparison without any systematic and consistent direction.

**Fig 4.**
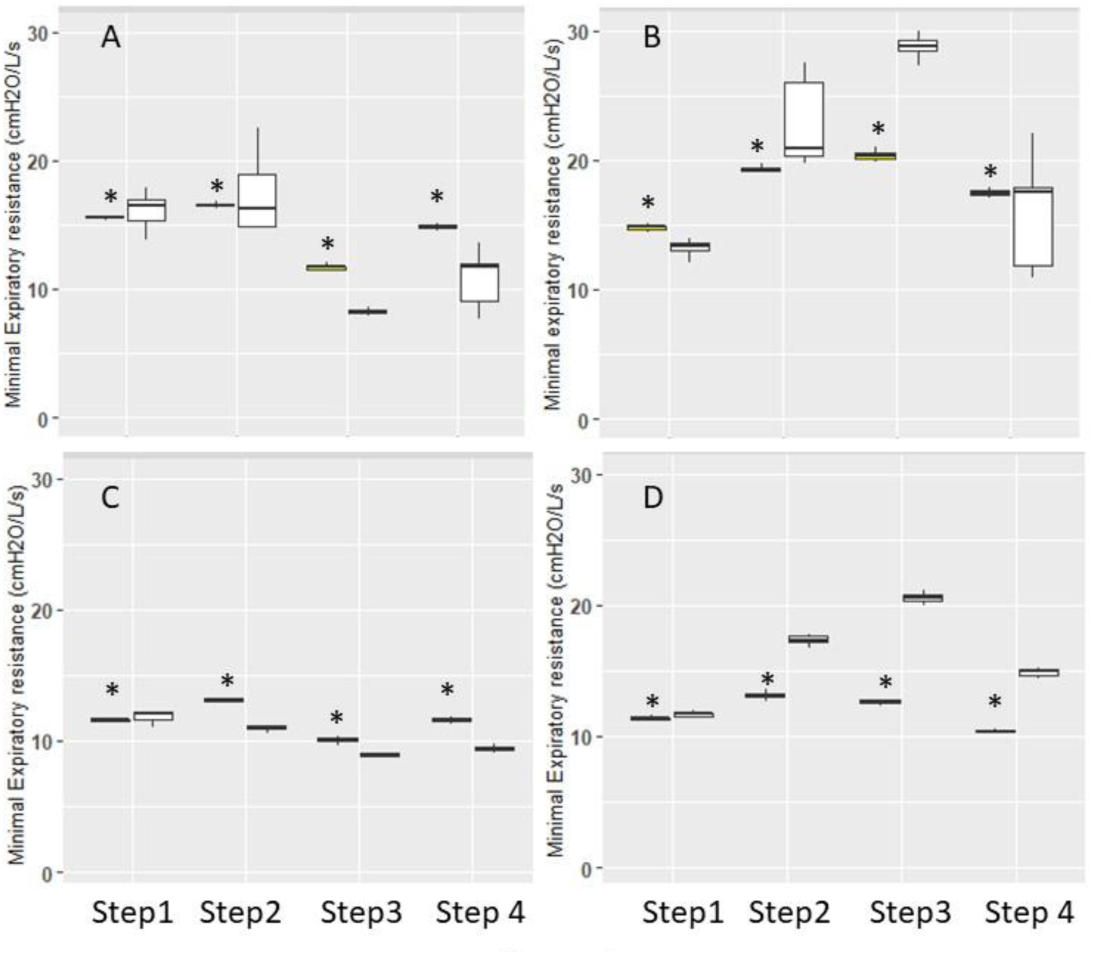
Expiratory resistance. Box-and-Whisker plots of minimal expiratory resistance in double (panel A and B) and triple circuits (panel C and D) with Elisée 350 (yellow) and V500 (white) ventilators during the different steps. Panels A and C are volume control and B and D pressure control. *P<0.05 vs. V500.

## Discussion

We found that: 1) the target V_T_ was achieved by the tested ventilators in volume and pressure control when they faced two symmetrical lungs, 2) asymmetrical C-R changed V_T_ distribution between patients, 3) the risk for air recirculation from one patient to the other was related to the difference in lung time constants and also to the ventilator, 4) the performance of the two ventilators was close.

The shortage in ventilators to support COVID-19 patients in acute respiratory failure results from the imbalance between an acute enormous demand and a limited supply. The response of the healthcare system was a dramatic increase in the number of ICU beds in a very short period but the number of ventilators available was an issue. Trying to share ventilators is typical behavior of the “we have to do something” concept [22] to provide a fair allocation of resources [23-25] in the COVID-19 pandemics.

Started before the current COVID-19 pandemic [26, 27], it was stressed that shared ventilation cannot support its use in mass causality because V_T_ was too much variable across C-R conditions and largely dependent on changes in C [27]. Since then, the current COVID-19 pandemic prompted additional bench studies to extend these previous results and proposed solutions to try to overwhelm some related issues [8, 11, 28-30] (Table 4).

**Table 4.** Summary of the bench studies on shared ventilation between two patients

| Author | Ventilator | Lung model | Data logger | VT target for ventilator (ml) | PEEP (cmH<sub>2</sub>O) | RC patient 1 | RC patient 2 | V<sub>T</sub> Volume control (ml) patients 1/2 | V<sub>T</sub> Pressure control (ml) patients 1/2 |
| --- | --- | --- | --- | --- | --- | --- | --- | --- | --- |
| Chatburn | Servo-i | ASL 5000 | ASL 5000 | 800 | 15 | 10-45 | 10-45 | 396/392 | 421/417 |
|  |  |  |  |  |  | 10-45 | 10-20 | 406/293 | 500/272 |
|  |  |  |  |  |  | 10-50 | 10-20 | 467/267 | 485/250 |
|  |  |  |  |  |  | 10-45 | 15-45 | 440/349 | 457/369 |
|  |  |  |  |  |  | 10-45 | 30-45 | 499/216 | 493/241 |
|  |  |  |  |  |  | 10-20 | 25-60 | 406/382 | 376/412 |
| Epstein | Servo air | NA | PF300 | 1000 | 8 | NA-37 | NA-24 | 473/314 | 475/333 |
| Herrmann | Servo 300 | DemoLung in one patient<br>Parabolic resistor and anesthesia reservoir bag in the other | Florian Monitor | 1000 | 5 | 20-50 | 20-50 | 438/482 | 443/475 |
|  |  |  |  |  |  | 20-50 | 20-22 | 592/337 | 460/196 |
|  |  |  |  |  |  | 20-50 | 20-7.5 | 735/159 | NA/NA |
|  |  |  |  |  |  | 20-50 | 20-35 | NA/NA | 451/332 |
| Han | PB840 | QuickLung | FlexMed Gr | 800 | 5 | NA-50 | NA-10 | NA/NA | 390/262 |
| Tonetti | Siaretron 4000 T (turbine) | Michigan Instruments 5601 | NA | 960 | 15 | 5-50 | 5-50 | NA/NA | 470/470 |
|  |  |  |  |  |  | 5-40 | 5-60 | NA/NA | 340/540 |
|  |  |  |  |  |  | 5-50 | 20-50 | NA/NA | 480/400 |

V<sub>T</sub>=tidal volume, PEEP=positive end-expiratory pressure, R=resistance, C=compliance, NA=not available

We found that pressure control should be the mode of choice because it preserves V_T_ in the least injured lung while volume control sets the healthier lung to overdistension and the worst lung to hypoventilation. By comparison to the step 1 the introduction of asymmetry in pressure control does not compromise the target V_T_ of the lung whose R-C set is not too much modified (lung A steps 2 and 4 in design 2 lungs, lung C all steps in design 3 lungs). In steps 2-4 adjusting the pressure preset to restore the targeted V_T_ in each lung will increase V_T_ in the preserved lung. Therefore, technical innovations have been proposed to individualize ventilator settings in each patient, such as set inspiratory pressure, PEEP, and F_I_O_2_. These innovations include a one-way flow control valve at inspiratory and expiratory limbs in each patient [12], a fixed pressure resistor regulator added at the inspiratory limb [31], a variable flow restrictor at the inspiratory limb, and a one-way valve at the expiratory limb [32], a flow restrictor on-way valve at the outlet of the ventilator [9], and bag-in-the box [29]. It should be noted that even though some of the interventions described above have been tested in a few patients [12], the experience is limited, they are complex to use and may generate further severe problems, as in case of an acute change in respiratory mechanics or gas exchange in one or two patients if the staff is not well trained.

The present study brings up new findings by testing two ventilators of different categories and measured air recirculation and expiratory resistance of the ventilator valve. Even though most of the differences in V_T_ between ventilators were statistically significant the clinical significance of them was irrelevant, meaning that the performance of the lower-level ventilator was close to that of the ICU ventilator. Therefore, the present findings suggest that the shortage of ICU-dedicated ventilators can be overcome by using safely lower-level ventilators like the one we tested.

The amount of rebreathing air from one patient to the other is another issue of shared ventilation. We quantified this in the present study both during inspiration and expiration (Figure 2). Even though the recirculating air is lower than the anatomical dead space (including the endotracheal tube volume) and arises at the end of the inspiration of the patient it would not induce CO_2_ retention but sets the patient at an increased risk of cross-infection. Rebreathing can be explained by a difference of “plateau pressure” between the two patients as depicted in figure 2. We expected that pressure control should prevent rebreathing from the following considerations. In terms of pressurization pressure control mode tends to favor fast and high pressurization finishing with a plateau while the volume control mode with its volume target tends to use less fast pressurization with a continuous increase of the pressure. In other words, we can expect that the pressure control mode is, at the end of the inspiration phase closer to an equilibrium situation for the plateau pressures than the volume control mode. The difference in rebreathing observed between the two ventilators is not simple to explain. Nevertheless, the difference observed in minimal expiratory resistance may also suggest that the opening speed of expiratory valves may be different between the ventilators. A delay to open the valve should favor gas recirculation between the two lungs at the start of the expiratory phase.

## Clinical implications

The clinical implication of present findings is that if one ventilator is dedicated to two or three patients, pressure control mode should be preferred and V_T_ and end-tidal CO_2_ of each patient should be closely monitored. Those patients should be managed by skilled caregivers, which would require an extensive education schedule in particular in those ICUs where pressure control is not routinely used. They would be able to understand and alert the clinical team of important changes in each of these patients.

An efficient filter upstream of the expiratory valve is mandatory. More complex implementations mentioned above have been achieved but may also generate problems. The recirculation of the air supposes not only some difference in pressure between the two patients but also an expiratory circuit common to the two patients (including the expiratory valve of the ventilator) sufficiently resistive. We recently found that expiratory resistance differs between ICU ventilators [21]. Therefore, adding a poorly resistive valve at the beginning of the expiratory circuit should deemphasize the phenomenon of air recirculation. Nevertheless, the presence of such a valve could affect the capacities of the ventilator to maintain the PEEP. Preclinical studies should be considered to increase our experience and knowledge in sharing ventilation in the dawn of the second wave of COVID-19 pandemic that is hitting the planet [33]

## Conclusion

The lower-level ventilator performed closely to the ICU-dedicated ventilator. Due to dependence of V_T_ to C pressure control should be used to maintain adequate V_T_ at least in one patient when C and/or R changes abruptly and monitoring of V_T_ should be done carefully.

## Data Availability

The data are available in the article as submitted.

## Acknowledgments

The authors would like to thank Michelin France for providing us with the flow-splitter, Maud Grammatica MD and Sylvain Guibert for their help to get the flow-splitter.

